# Clinical severity in Parkinson’s disease is determined by decline in cortical compensation

**DOI:** 10.1101/2023.04.16.23288636

**Authors:** Martin E. Johansson, Ivan Toni, Roy P.C. Kessels, Bastiaan R. Bloem, Rick C. Helmich

**Affiliations:** Radboud University Medical Center, Donders Institute for Brain, Cognition and Behaviour, Centre of Expertise for Parkinson & Movement Disorders, 6525 EN Nijmegen, the Netherlands; Radboud University, Donders Institute for Brain, Cognition and Behaviour, 6525 EN Nijmegen, the Netherlands; Radboud University Medical Center, Department of Medical Psychology, 6525 GA Nijmegen, The Netherlands; Radboud University Medical Center, Radboudumc Alzheimer Center, 6525 GA Nijmegen, The Netherlands; Vincent van Gogh Institute for Psychiatry, 5803 AC Venray, The Netherlands

**Keywords:** Parkinson’s disease, action selection, fMRI, compensation, inter-individual differences

## Abstract

Dopaminergic dysfunction in the basal ganglia, particularly in the post-commissural putamen, is often viewed as the primary pathological mechanism behind motor slowing (i.e., bradykinesia) in Parkinson’s disease. However, striatal dopamine loss fails to account for inter-individual differences in motor phenotype and rate of decline, implying that the expression of motor symptoms depends on additional mechanisms, some of which may be compensatory in nature. Building on observations of increased motor-related activity in the parieto-premotor cortex of Parkinson patients, we tested the hypothesis that inter-individual differences in clinical severity are determined by compensatory cortical mechanisms, and not just by basal ganglia dysfunction.

Using functional MRI, we measured variability in motor- and selection-related brain activity during a visuomotor task in 353 patients with Parkinson’s disease (≤5 years disease duration) and 60 healthy controls. In this task, we manipulated action selection demand by varying the number of possible actions that individuals could choose from. Clinical variability was characterized in two ways. First, patients were categorized into three previously validated, discrete clinical subtypes: diffuse-malignant (n=42), intermediate (n=128), or mild motor-predominant (n=150). Second, we used the total bradykinesia score across the entire sample as a continuous measure.

Patients showed motor slowing (longer response times) and reduced motor-related activity in the basal ganglia compared to controls. However, basal ganglia activity did not differ between clinical subtypes and was not associated with clinical bradykinesia scores. This indicates a limited role for striatal dysfunction in shaping inter-individual differences in symptom severity. Consistent with our hypothesis, we observed enhanced action selection-related activity in the parieto-premotor cortex of patients with a mild-motor predominant subtype, both compared to patients with a diffuse-malignant subtype and to controls. Furthermore, parieto-premotor activity was inversely related to bradykinesia, which points to a compensatory role.

We conclude that parieto-premotor compensation, rather than basal ganglia dysfunction, shapes inter-individual variability in symptom severity in Parkinson’s disease. Future interventions may focus on maintaining and enhancing compensatory cortical mechanisms, rather than only attempting to normalize basal ganglia dysfunction.

## Introduction

Bradykinesia is one of the cardinal motor symptoms of Parkinson’s disease and manifests as slowness during the selection and execution of voluntary movements.^1, 2^ The severity of bradykinesia, which varies substantially between individuals, has been considered an outcome of dopamine depletion and basal ganglia dysfunction.^3, 4^ However, motor symptoms may also be shaped by compensatory cortical processes.^2, 5–8^ It remains unclear to what extent such compensatory processes, along with inter-individual differences in their efficacy, contribute to clinical heterogeneity in Parkinson’s disease, over and above basal ganglia dysfunction. We tested this by analysing data from a cohort of patients (Personalized Parkinson Project;^9^ *N*=353) who performed an action selection task that is sensitive to bradykinesia whilst undergoing functional MRI.

In Parkinson’s disease, progressive degeneration of dopaminergic neurons in the substantia nigra leads to dopamine depletion in the striatum, resulting in dysfunctional basal ganglia output and impaired motor performance.^4, 10–14^ These motor impairments become particularly pronounced in situations where patients are required to make a voluntary choice between multiple competing movement options.^15–19^ However, striatal dopamine depletion occurs gradually over several years prior to the onset of bradykinesia.^7, 12, 20–24^ Furthermore, motor symptoms progress despite the fact that the motor region of the striatum (posterior putamen) is almost entirely depleted of dopamine four years after diagnosis.^20^ Additionally, several studies have failed to demonstrate an association between changes in motor symptom severity and striatal dopamine levels over time^12, 25–28^ (although see^29^). These observations strongly imply that basal ganglia dysfunction is not the sole mechanism underlying bradykinesia and action selection deficits in Parkinson’s disease.

Over the last decades, several studies have suggested that action selection may involve compensatory cerebral processes in Parkinson’s disease.^6^ Cerebral compensation has been conceptualized as a performance-enhancing recruitment of neural resources that enable individuals to meet moderately high task demands despite deficits in the neural mechanisms that typically support task performance.^30–32^ Such compensatory mechanisms are thought to be instantiated during the long pre-clinical phase of Parkinson’s disease, which may last for several years,^33, 34^ to stabilize behavioural performance as basal ganglia dysfunction gradually worsens.^5, 7, 35–37^ However, with disease progression, the degree of basal ganglia dysfunction will eventually exceed the capacities of compensatory mechanisms, leading to the appearance and subsequent worsening of motor deficits.^35, 38, 39^ Importantly, the efficacy of these compensatory mechanisms likely differ between individuals owing to idiosyncrasies in patterns of pathology (e.g. focal vs. diffuse propagation of α-synucleinopathy), and may therefore contribute to the clinical heterogeneity that characterizes Parkinson’s disease.^40, 41^

Demonstrating compensation with neuroimaging is not straightforward, since increased brain activity during a task may reflect either recruitment of compensatory resources or reduced efficiency of processes that support task performance.^31, 42^ Two basic functional criteria for establishing compensation have been suggested. First, it should be clear what is being compensated for, such as basal ganglia dysfunction in the case of Parkinson’s disease.^4^ Second, compensatory brain activity should have beneficial effects on behavioural performance.^30^ More generally, demonstrating compensation requires statistical power adequate to detect biologically plausible effects linking neural compensation and behavioural performance in Parkinson’s disease.^43, 44^ Recent meta-analyses of functional MRI studies have shown that patients with Parkinson’s disease have increased motor-related activation in parieto-premotor regions^13, 14^. This cortical activity may be compensatory, e.g. enhancing goal-directed control during motor execution, but most studies had sample sizes inadequate to quantify brain-behaviour correlations. Other studies have focused on pre-symptomatic carriers of gene mutations associated with a high risk of developing Parkinson’s disease, who showed intact behavioural performance in combination with increased premotor activity relative to healthy controls during action selection.^45, 46^ This selection-related increase in premotor activation may be compensatory in the pre-symptomatic stage, but it decreases as motor symptoms worsen after the onset of Parkinson’s disease.^47^ Finally, we and others have shown that patients with Parkinson’s disease rely more heavily on the extrastriate visual cortex during motor imagery of their most affected hand,^48^ and that disruption of this region with transcranial magnetic stimulation impaired motor imagery in Parkinson’s disease patients, but not in healthy controls.^49^ This imagery-related extrastriate visual cortex activity might be compensatory, but a relationship with actual motor behaviour has not yet been established.

Here, we tested the hypothesis that the clinical heterogeneity in Parkinson’s disease depends on compensatory parieto-premotor function, over and above basal ganglia dysfunction. We used a motor task in combination with functional MRI to assess motor- and selection-related brain activity in early-to-moderate^50^ Parkinson’s disease patients. We verified that Parkinson’s disease is associated with reduced activity in the basal ganglia by comparing task-related activity in patients against healthy controls. We also tested for normalizing effects of dopamine replacement therapy on motor network dysfunction. The novelty of this study concerns the neural mechanisms underlying clinical heterogeneity in Parkinson’s disease. First, we compared clinical subtypes (mild-motor predominant, intermediate, and diffuse-malignant) that were defined based on motor symptoms, cognitive performance, REM sleep behaviour disorder, and autonomic dysfunction.^51^ Second, we quantified the relationship between inter-individual variability in brain activity and the clinically rated severity of bradykinesia. Our findings show that enhanced parieto-premotor activity is related to a more benign subtype of Parkinson’s disease and to less severe bradykinesia, suggesting that it may be compensating for a basal ganglia deficit.

## Materials and methods

### Participants

Data from 367 patients diagnosed with idiopathic Parkinson’s disease and 60 healthy controls were retrieved from the Personalized Parkinson Project database in March 2022. The Personalized Parkinson Project is an ongoing single-centre longitudinal cohort study taking place at Radboud University Medical Center (Nijmegen, the Netherlands; ClinicalTrials.gov identifier: NCT03364894 and NCT05169827).^9^. All patients underwent sequential motor symptom assessments off (i.e., 12 hours since the last dose of dopaminergic medication) and on dopaminergic medication with the Movement Disorders Society Unified Parkinson Disease Rating Scale part III (MDS-UPDRS III).^52^ MRI measurements were acquired in the on-medicated state. 56 patients returned for identical MRI measurements off medication. Half of these 56 patients were assessed within 3 months after their first on-state measurement. The other half were assessed within three months prior to a two-year follow-up visit, to reduce potential learning effects and even out the distribution of disease durations in the off-medicated group. Healthy controls were matched to the off-medicated group with respect to age, sex, and handedness. Written informed consent was obtained for all participants in accordance with the Declaration of Helsinki. The study was approved by a medical ethical committee (METC Oost-Nederland, formerly CMO Arnhem-Nijmegen; #2016-2934 and #2018-4785). See Appendix 1 for detailed information on inclusion and exclusion criteria. During baseline assessments, diagnoses of eight patients were re-evaluated to either a form of atypical parkinsonism (*N*=2) or other (*N*=6). Diagnosis re-evaluations at two-year follow-up confirmed that an additional six patients did not have Parkinson’s disease (three multiple system atrophy, two progressive supranuclear palsy, and one indeterminate). All patients with a verified non-Parkinson’s disease diagnosis at either baseline or follow-up were excluded from further analysis, resulting in a total sample size of 353 on-medicated patients (of whom 55 also had an off-medication assessment) and 60 controls. Demographic information can be found in Table 1.

**Table 1.**
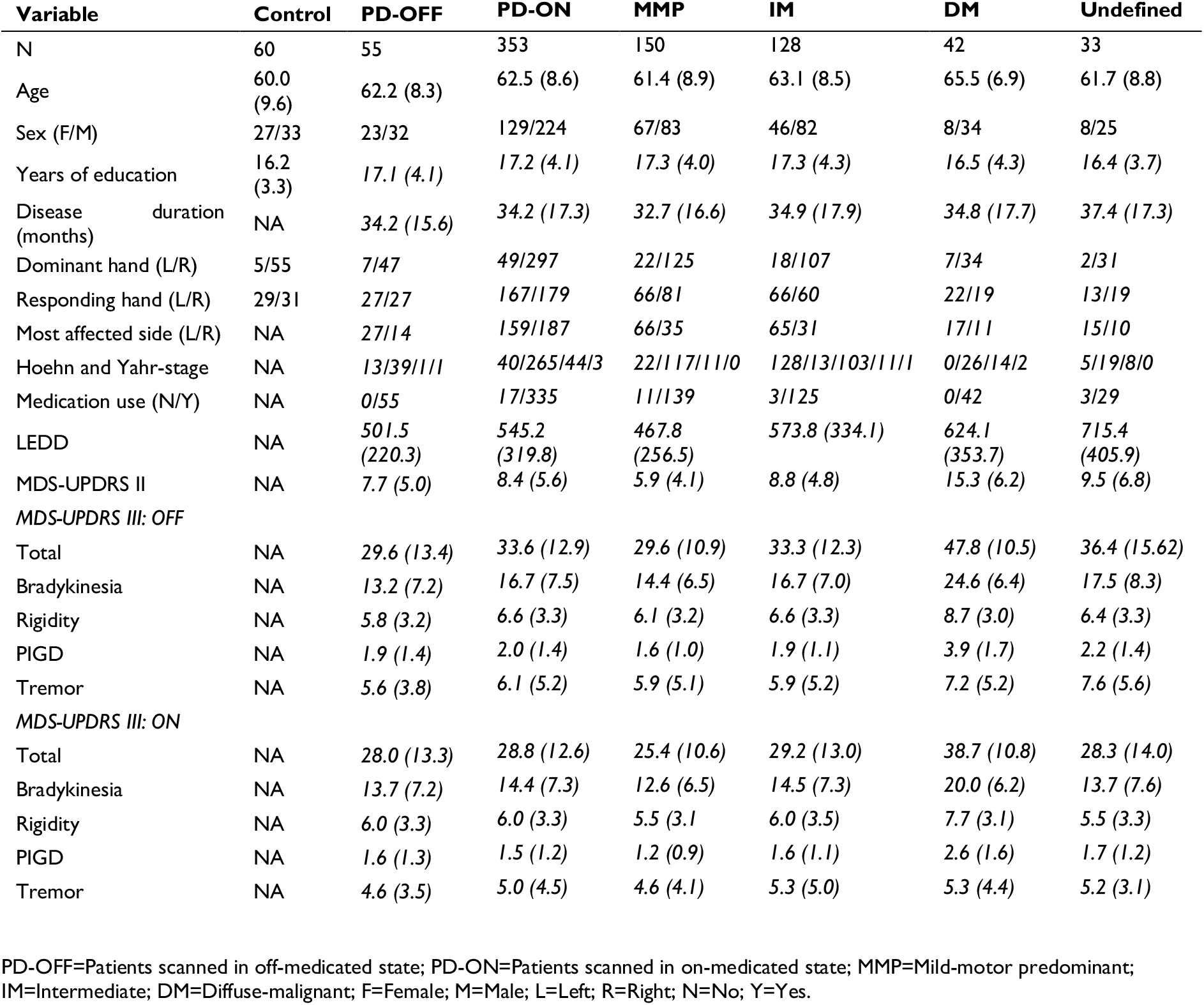
Demographic information and baseline characteristics

### Clinical measurement of bradykinesia

A subscore for bradykinesia symptoms was defined by summing together 11 items of the MDS-UPDRS III that assesses bradykinesia (4-9 and 14).^53, 54^ Bradykinesia scores acquired in the off-medicated state were used to test for associations with task performance and brain activity.

### Subtyping

We utilized a recently developed^51^ and previously validated^41, 55^ clinical subtype classification to parse clinical heterogeneity. The classification used in the present study has been described in detail in a previous publication where we investigated clinical differences between subtypes in the larger Personalized Parkinson Project cohort.^41^ In short, patients were classified based on motor symptoms, cognitive function, REM movement sleep behaviour disorder, and autonomic function (see Appendix 1). Classification resulted in three distinct subtypes: diffuse-malignant (n=42), intermediate (n=128), and mild-motor predominant (n=150). 33 patients could not be classified due to missing data. In short, the diffuse-malignant subtype showed relatively severe motor symptoms, impairment in more clinical domains, and faster progression in comparison to the mild-motor predominant subtype.^41^

### Action selection task

#### Task instructions

Participants performed an action selection task whilst undergoing mixed block/event-related functional MRI (Fig. 1).^56–58^ This task was specifically designed to target Parkinson’s disease-related deficits in voluntary action selection that are thought to underlie bradykinesia.^18, 19^ Participants were instructed to respond to highlighted cues with a single button press as quickly and as accurately as possible, and to try to make equal use of all response options. The number of highlighted cues varied between one and three choices. If multiple cues were highlighted, participants were instructed to choose and respond to one cue only. See Appendix 1 for a more detailed account of the task.

**Fig. 1.**
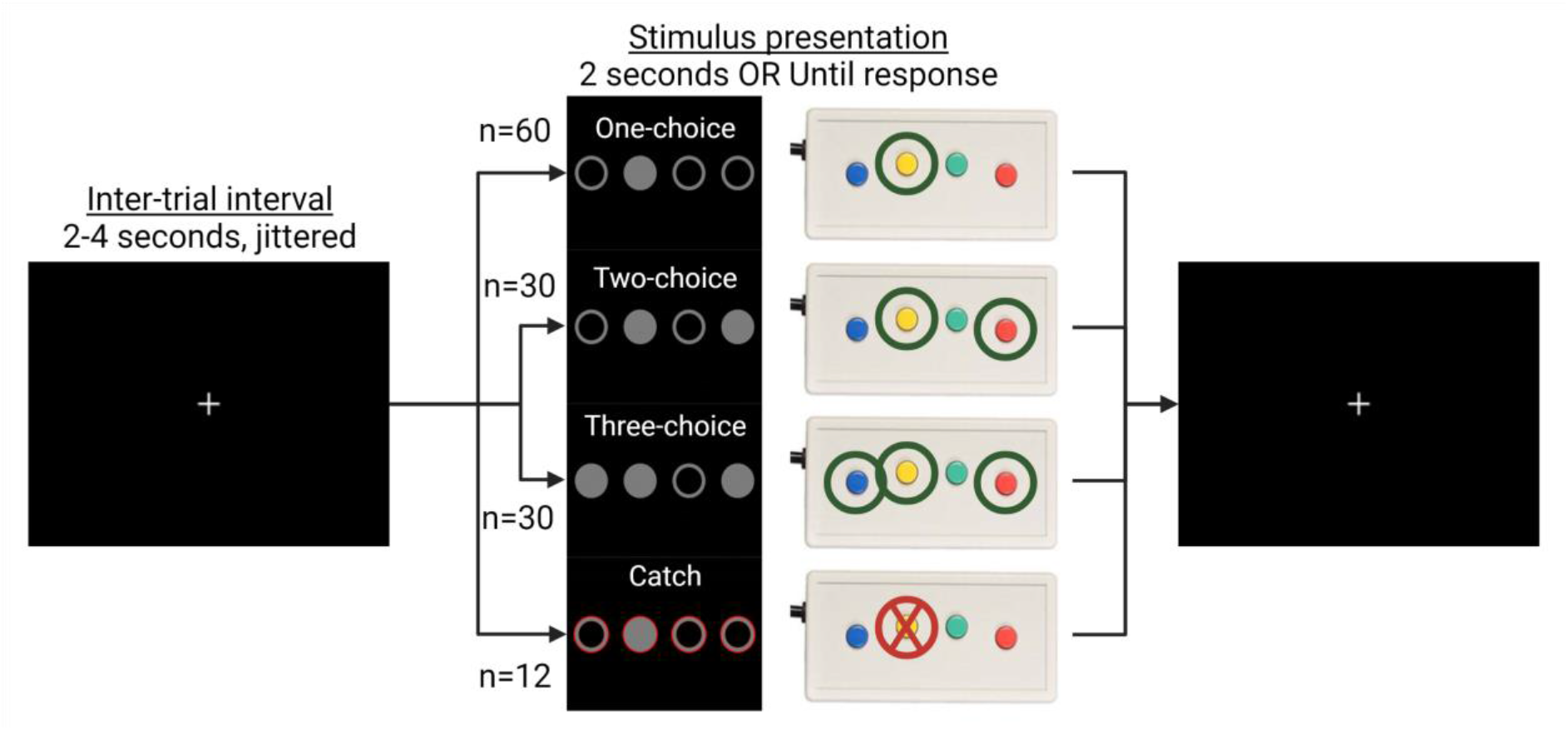
Action selection task. Participants respond to highlighted circles with one out of four response buttons. When multiple circles are highlighted, participants are instructed to select one response button. Action selection demand is parametrically manipulated by varying the number of highlighted circles that are presented. During catch trials, participants are instructed to withhold a response. No feedback was given to indicate the correctness of responses.

#### Measurements of behavioural performance

Response times and error rates were aggregated within participants by trial condition (one-choice, two-choice, three-choice) and block (one, two, three), resulting in nine values per participant for each measurement. Response times were aggregated by taking the median after excluding misses, incorrect responses, and response times below 300 milliseconds. Error rates were similarly aggregated by taking the number of incorrect trials divided by the total number of correct and incorrect trials after excluding misses. In trials where multiple choices were possible, participants sometimes had the option to either repeat their previous response or switch to a new response. Based on such trials, two additional metrics (response variability and switching) were calculated to characterize the use of stereotyped response strategies.^47^ First, for each of the four response options, the number of button presses were calculated. Response variability (i.e., the degree to which all available response options were used) was subsequently characterized as a coefficient of variation, calculated by taking the standard deviation of button presses across response options divided by the mean. Larger coefficients of variation indicate that button presses are not uniformly distributed. Second, response switching (i.e., the degree to which the same response was repeated over consecutive trials) was characterized as the ratio of switches relative to the total number of switches and repeats. Larger ratios indicate that response repetitions are more likely than switches.

### Image acquisition

All scans were acquired using a Siemens MAGNETOM Prisma 3T (Siemens, Erlangen, Germany) equipped with a 32-channel head coil. T1-weighted anatomical images were acquired using a magnetization-prepared rapid gradient-echo sequence (TR/TE/TI=2000/2/880 milliseconds; flip angle=8°; voxel size=1.0×1.0×1.0 millimetres; slices=192; FOV=256 millimetres; scanning time=5 minutes). T2*-weighted functional images were acquired during the performance of the action selection task using a multi-band sequence (TR/TE=1000/34 milliseconds; acceleration factor=6; acquisition mode=interleaved; flip angle=60°; voxel-size=2.0×2.0×2.0 millimetres; slices=72; FOV=210 millimetres; scanning time=9-10 minutes).

### Image acquisition and pre-processing

Pre-processing of fMRI data was performed using a standardized pipeline in *fmriprep* (v20.2.1).^59^ In short, functional images were motion- and slice time-corrected, and normalized to MNI152Lin6Asym-space. Lastly, corrected and normalized images were spatially smoothed with Gaussian kernel of 6 millimetres at full-width half-maximum. See Appendix 1 for detailed information.

### First-level analysis

SPM12 (https://www.fil.ion.ucl.ac.uk/spm/software/spm12) was used to carry out all first-level and group-level analyses. Task regressors were generated for one-, two-, and three-choice conditions by convolving cue onsets with a canonical hemodynamic response function. Cue duration was defined as the average response time across choice conditions. Time derivatives were included for each condition together with parametric regressors for response time and their first-order derivatives. Additional regressors were generated for catch trials and incorrect responses. Confound time series were included in the first-level model to correct for anatomical and motion-related sources of noise (see Appendix 1). Task-related activity for each choice condition was defined by contrasting each separate task regressor against an implicit baseline (one-choice>0; two-choice>0; three-choice>0; catch>0). The resulting contrast images were used as inputs for group comparisons. Additional contrasts were formed to encode motor-related ([one-choice, two-choice, three-choice]>0) and selection-related (two-choice>one-choice; three-choice>one-choice) activity (see Supp. Fig. 1 for mean activation associated with each contrast of interest). These contrasts were used to assess associations with bradykinesia severity. Contrast images of all participants who responded with the left side were flipped horizontally to ensure that the most-affected side was consistent across patients.

### Statistical analysis

#### Behavioural performance

The influences of disease status (between-subjects factor GROUP: patient vs control), dopaminergic medication (within-subjects factor GROUP: on vs off medication), and subtype (between-subjects factor GROUP: mild-motor predominant vs intermediate vs diffuse-malignant) were assessed with linear mixed-effects models for log-transformed response times and with binomial logistic mixed-effects models for error rates (weighted by total number of responses) using the lme4-package^60^ in R 4.2.1 (R Core Team, 2022). Each model included fixed effects for GROUP and CHOICE (within-subjects factor CHOICE: one-choice, two-choice, three-choice) as well as their interaction. Repeated measures were accounted for with by-subject random intercepts. By-block random intercepts were included to account for task habituation. Age, sex, and years of education were included as additional covariates of non-interest. Associations between behavioural performance and bradykinesia severity were assessed within patients using the same model formula, with the exception that the factor of GROUP was removed and a term was added for SYMPTOM SEVERITY, defined as the bradykinesia subscore of the MDS-UPDRS III assessed in an off-medicated state. Models were fitted using a restricted maximum likelihood approach. *P*-values for fixed effects were derived through type III analyses of deviance using Wald *χ*^2^ tests. Response variability and switching were analysed using one-way ANCOVAs, with GROUP as a between-subjects factor and age, sex, and years of education as covariates of non-interest. Two-tailed post hoc t-tests were performed on estimated marginal means. Participants with less than 25% correct responses on one-choice trials, averaged across blocks, were excluded from further analysis. In analyses of response variability and switching, additional outliers scoring above or below three standard deviations from the mean were excluded.

#### Brain activity

Group comparisons of task-related brain activity closely followed the group comparisons that were conducted for behavioural performance. Three separate repeated-measures ANCOVAs, implemented using the full factorial design option in SPM12, were conducted to test for effects of disease status, medication, and subtype on brain activity. Estimates of brain activity for each task condition were used as inputs. Contrasts were set up to compare motor- and selection-related activity between groups. The comparison between subtypes was followed by a post hoc comparison between each subtype and controls. One-way ANCOVAs with the bradykinesia subscore as a regressor of interest were used to test the association between brain activity and symptom severity, and were fitted against first-level contrasts of motor- and selection-related activity. Mean framewise displacement, age, and sex were included as covariates of non-interest. Cluster-based thresholding, with a cluster-forming threshold of *Z*=3.1, was used to correct for family-wise error at *P*<0.05.^61^ See Appendix 2 for average task-related effects on brain activity. Anatomical labels and functional subdivisions of significant clusters were derived from the JuBrain Anatomy Toolbox (v3.0)^62^ and Glasser atlas,^63^ respectively.

### Data availability

The data that support the findings of this study are available upon request only, to ensure the privacy of the participants. A data acquisition request can be sent to the corresponding author. All analysis code used in the present study is freely available at https://github.com/mejoh/Personalized-Parkinson-Project-Motor.

## Results

### The influence of disease status

#### Behavioural performance

Response times were *longer* in Parkinson patients than controls (Fig. 2A; main effect of GROUP [*χ*^2^(1)=15.2, *P*<0.001, *η^2^_p_*=0.04]; patient>control [*log-ratio*=1.08, *SE*=0.02, *t-ratio*(391)=3.9, *P*<0.001]), and they increased with action selection demand (main effect of CHOICE [*χ*^2^(2)=80.4, *P*<0.001, *η*^2^ =0.07]; intermediate>low [*log-ratio*=1.08, *SE*=0.006, *t-ratio*(3162)=14.0, *P*<0.001], high>low [*log-ratio*=1.08, *SE*=0.006, *t-ratio*(3162)=13.6, *P*<0.001]). Patients were not disproportionally slower for multi-choice trials than controls (no GROUP×CHOICE interaction, *P*=0.86).

**Fig. 2.**
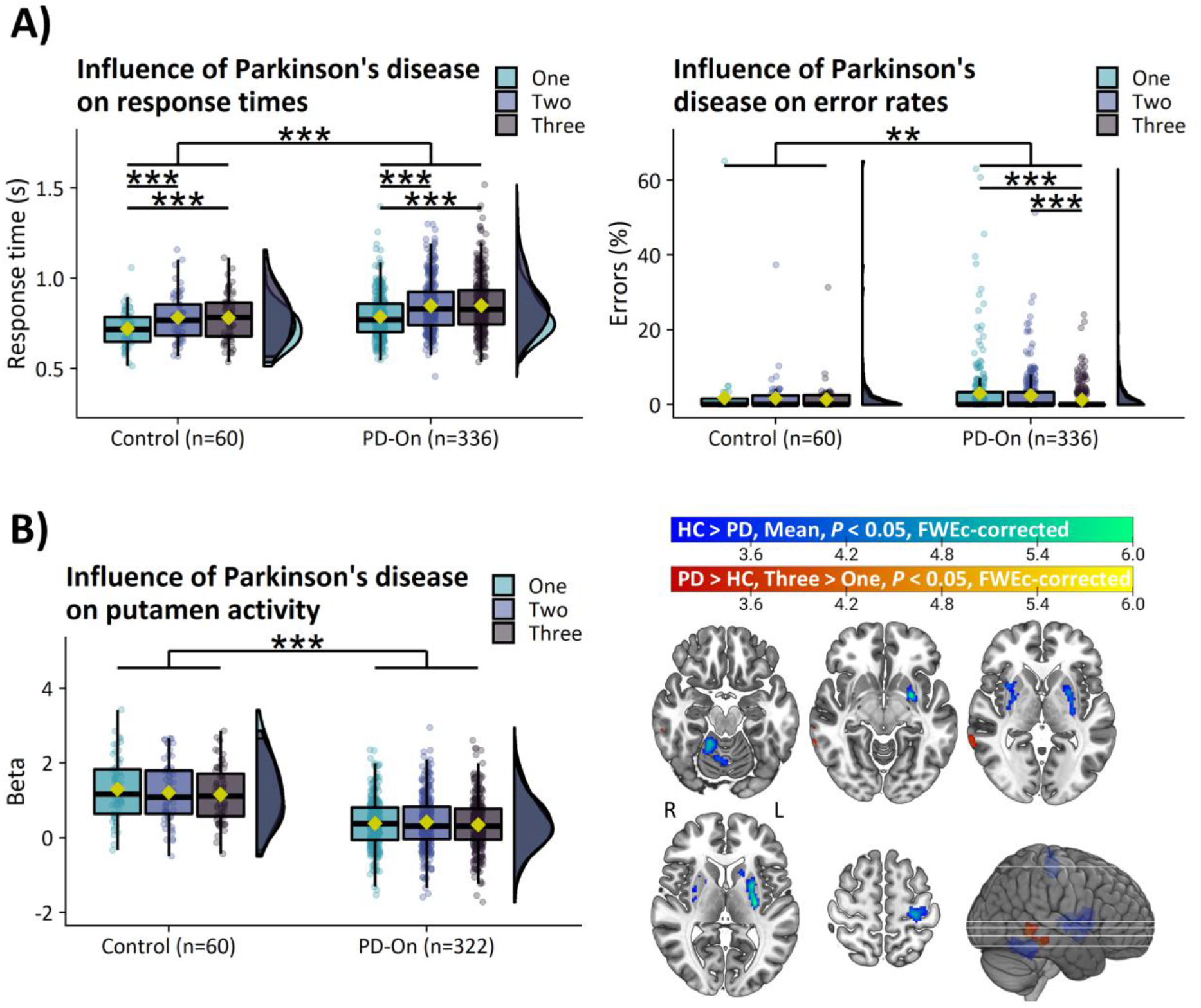
Action selection deficits and their relation to bradykinesia. (**A**) Parkinson’s disease leads to general deficits in task performance. (**B**) Parkinson’s disease leads to reduced motor-related activity in a network of core motor regions and reduced selection-related deactivation in the middle temporal gyrus. HC=Healthy control, PD-On=On-medicated Parkinson’s disease, s=Seconds, SE=Standard error, OR=Odds ratio, *=*P*<0.05, **=*P*<0.01, ***=*P*<0.001, FWEc=Family-wise error cluster. Box-plots show the first and third quartiles (boxes), median (horizontal black line), mean (yellow rectangle), and 1.5×interquartile range (whiskers). Brain images show *T*-values of significant clusters.

Error rates were *higher* in Parkinson patients than controls (Fig. 2A; main effect of GROUP [*χ*^2^(1)=7.6, *P*=0.006]; patient>control [*OR*=1.6, *SE*=0.4, *Z-ratio*=1.9, *P*=0.052]). Patients tended to make more errors specifically during single-choice versus multi-choice trials compared to controls, but this effect did not reach significance (GROUP×CHOICE [*χ*^2^(2)=5.8, *P*=0.055]; one-choice>three-choice, patient>control [*OR*=1.9, *SE*=0.5, *Z*-*ratio*=2.4, *P*=0.049]).

There was no effect of GROUP on response variability (Supp. Fig. 2A; *P*=0.09) or switching (Supp. Fig. 2B; *P*=0.19).

#### Brain activity

**Motor-related activity:** patients with Parkinson’s disease showed *reduced* motor-related activity in the bilateral putamen, right cerebellar lobule IV-V, and left precentral gyrus (area 4) compared to controls (Table 2; Fig. 2B; control>patient, mean activity>baseline).

**Table 2.** Statistically significant clusters from voxel-wise comparisons between groups

| Anatomical label<br>(% cluster volume<br>in area) | Area | P-value<br>(FWEc-<br>corrected) | Cluster extent<br>(voxels) | Max t-value | MNI:<br>X, Y, Z |
| --- | --- | --- | --- | --- | --- |
| **Influence of Parkinson's disease** |  |  |  |  |  |
| **PD-ON vs. Control** |  |  |  |  |  |
| **Control > PD-ON, Mean > Baseline** |  |  |  |  |  |
| L Putamen (78%) |  | <0.001 | 571 | 6.1 | -25,3,4 |
| R Cerebellum (64%) | IV-V | <0.001 | 383 | 5.5 | 17,-45,-20 |
| L PreCG (70%) | 4 | <0.001 | 423 | 5.4 | -23,-25,66 |
| R Putamen (85%) |  | <0.001 | 217 | 4.4 | 29,-3,2 |
| **PD-ON > Control, Three > One** |  |  |  |  |  |
| R MTG (89%) | TEIp | 0.011 | 156 | 5.2 | 61,-37,-14 |
| **Influence of medication** |  |  |  |  |  |
| **PD-ON vs. PD-OFF** |  |  |  |  |  |
| Ns. |  |  |  |  |  |
| **Influence of subtype** |  |  |  |  |  |
| **MMP vs. IM** |  |  |  |  |  |
| Ns. |  |  |  |  |  |
| **MMP vs. DM** |  |  |  |  |  |
| **MMP > DM, Mean > Baseline** |  |  |  |  |  |
| R PostCG (34%) | 2 | 0.003 | 166 | 4.0 | 51,-19,38 |
| **MMP > DM, Three > One** |  |  |  |  |  |
| R MFG (48%) | i6-8 | <0.001 | 401 | 4.6 | 33,17,52 |
| R IPL(57%) | PGi | 0.042 | 99 | 4.0 | 47,-55,38 |
| R SPL (33%) | 7Pm | 0.021 | 116 | 3.9 | 9,-71,54 |
| **IM vs. DM** |  |  |  |  |  |
| **IM > DM, Three > One** |  |  |  |  |  |
| L SFG (47%) | 6a | 0.016 | 123 | 4.7 | -21,3,56 |
| **Post hoc comparison between subtypes and controls** |  |  |  |  |  |
| **MMP vs. Control** |  |  |  |  |  |
| **MMP > Control, Three > One** |  |  |  |  |  |
| R IPL (43%) | PFm | <0.001 | 214 | 5.5 | 51,-63,46 |
| R MTG (40%) | TEIp | 0.001 | 185 | 4.5 | 61,-37,-14 |
| R MFG (54%) | 8Av | 0.015 | 122 | 4.5 | 27,11,40 |
| R SFG (61%) | 9a | 0.031 | 104 | 4.0 | 23,58,30 |
| **IM vs. Control** |  |  |  |  |  |
| Ns. |  |  |  |  |  |
| **DM vs. Control** |  |  |  |  |  |
| **Control > DM, Three > One** |  |  |  |  |  |
| L SFG (49%) | 6a | <0.001 | 226 | 4.8 | -25,5,60 |

Ns.=No significant clusters; PD-ON=Patients scanned in on-medicated state; MMP=Mild-motor predominant; IM=Intermediate; DM=Diffuse-malignant; PreCG=Precentral gyrus; MTG=Middle temporal gyrus; PostCG=Postcentral gyrus; MFG=Middle frontal gyrus; IPL=Inferior parietal lobule; SPL=Superior parietal lobule; SFG=Superior frontal gyrus; FWEc=Family-wise error cluster. Anatomical labels were derived from the Anatomy Toolbox v2.2. Area labels were derived from the Glasser atlas.

**Selection-related activity:** patients with Parkinson’s disease showed *increased* selection-related activity in the right middle temporal gyrus (TE1p) compared to controls (Table 2; Fig. 2B; patient>control, three-choice>one-choice). This increase resulted from reduced task-related deactivation in patients compared to controls (Supp. Fig. 3).

### The influence of clinical subtype

#### Behavioural performance

The influence of action selection demand on response times differed between subtypes (Fig. 3A; GROUP×CHOICE [*χ*^2^(4)=11.7, *P*=0.02, *η^2^_p_*=0.005]). The difference in response times between high and low action selection demand was *increased* for intermediate compared to mild-motor predominant patients (intermediate>mild-motor predominant, three-choice>one-choice [*log-ratio*=1.027, *SE*=0.01, *t-ratio*(2448)=2.8, *P*=0.032]), but not for diffuse-malignant compared to mild-motor predominant patients (*P*=0.99).

**Fig. 3.**
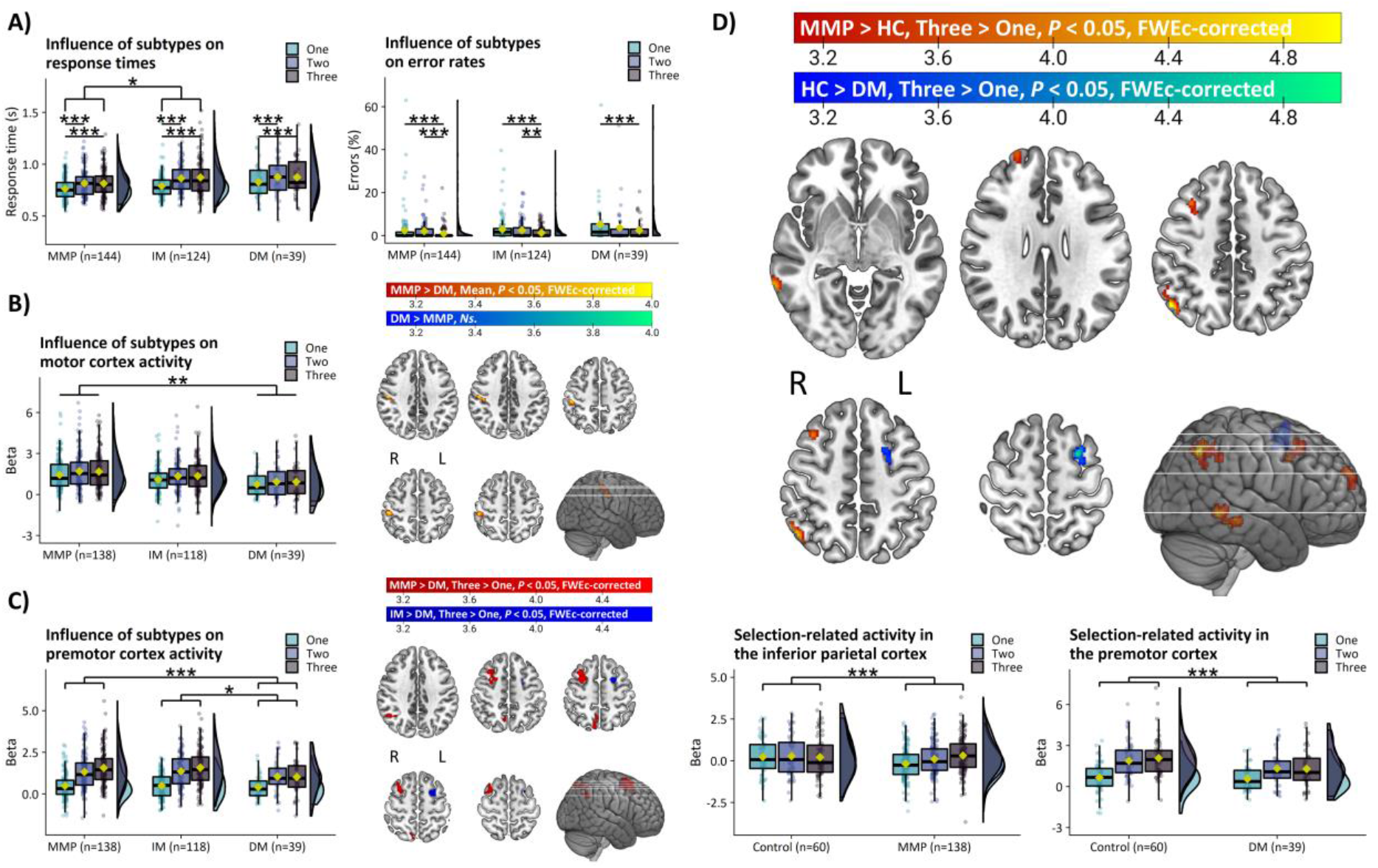
Evidence for parieto-premotor compensation. (**A**) Greater selection-related increase in response times in intermediate compared to mild-motor predominant patients. (**B**) Reduced motor-related activity in the motor cortex in diffuse-malignant compared to mild-motor predominant patients. (**C**) Reduced selection-related activity in diffuse-malignant compared to mild-motor predominant and intermediate patients. (**D**) In comparison to controls, mild-motor predominant patients show increased selection-related parieto-premotor activity whereas diffuse-malignant patients show the opposite. s=Seconds, MMP=Mild-motor predominant, IM=Intermediate, DM=Diffuse-malignant, *=*P*<0.05, **=*P*<0.01, ***=*P*<0.001, FWEc=Family-wise error cluster. Box-plots show the first and third quartiles (boxes), median (horizontal black line), mean (yellow rectangle), and 1.5×interquartile range (whiskers). Brain images show *T*-values of significant clusters.

Error rate increased as a function of action selection demand (Fig. 3A; main effect of CHOICE [*χ*^2^(2)=34.6, *P*<0.001]; low>intermediate [*OR*=1.2, *SE*=0.11, *Z-ratio*=2.4, *P*=0.044], low>high [*OR*=2.5, *SE*=0.28, *Z-ratio*=8.1, *P*<0.001], intermediate>high [*OR*=2.0, *SE*=0.26, *Z-ratio*=5.5, *P*<0.001]).

There was no effect of GROUP on response variability (Supp. Fig. 2C; *P*=0.41) or switching (Supp. Fig. 2D; *P*=0.54).

#### Brain activity

**Motor-related activity:** Parkinson patients with a mild-motor predominant subtype had *increased* motor-related activity in the right postcentral gyrus (area 2) compared to patients with a diffuse-malignant subtype (Table 2; Fig. 3B; mild-motor predominant>diffuse-malignant, mean activity>baseline). There were no differences in basal ganglia activity between subtypes (Supp. Fig. 4).

**Selection-related activity:** Patients with a mild-motor predominant subtype had *increased* selection-related activity in the right middle frontal gyrus (i6-8), right inferior parietal lobule (PGi), and right superior parietal lobule (7Pm) compared to patients with a diffuse-malignant subtype (Table 2; Fig. 3C; mild-motor predominant>diffuse-malignant, three-choice>one-choice). Furthermore, patients with an intermediate subtype also had *increased* selection-related activity in the left superior frontal gyrus (6a) compared to the diffuse-malignant subtype (Fig. 3C; intermediate>diffuse-malignant, three-choice>one-choice). These results suggest that a more benign clinical phenotype is associated with *higher* activation in a network that involves premotor, inferior parietal, and superior parietal cortex.

Post hoc comparisons of selection-related activity between subtypes and controls were carried out to assess the clinical relevance of the results above. Patients with a mild-motor predominant subtype had *increased* selection-related activity in the right inferior parietal lobule (PFm), right middle temporal gyrus (TE1p), right middle frontal gyrus (8Av), and right superior frontal gyrus (9a) compared to controls (Fig. 3D; mild-motor predominant>control, three-choice>one-choice). In contrast, patients with a diffuse-malignant subtype had *decreased* selection-related activity in the left superior frontal gyrus (6a) compared to controls (Fig. 3D; control>mild-motor predominant, three-choice>one-choice).

### Associations with bradykinesia severity

#### Behavioural performance

Response times *increased* as a function of bradykinesia severity (Fig. 4A; main effect of SEVERITY [*χ*^2^(1)=)=12.5, *P*<0.001, *η*^2^ =0.03]) and action selection demand (main effect of CHOICE [*χ*^2^(2)=380, *P*<0.001, *η^2^_p_*=0.12]; intermediate>low [*log-ratio*=1.075, *SE*=0.005, *t-ratio*(2666)=17.2, *P*<0.001], high>low [*log-ratio*=1.073, *SE*=0.005, *t-ratio*(2666)=16.6, *P*<0.001]). There was no interaction between SEVERITY and CHOICE (*P*=0.64).

**Fig. 4.**
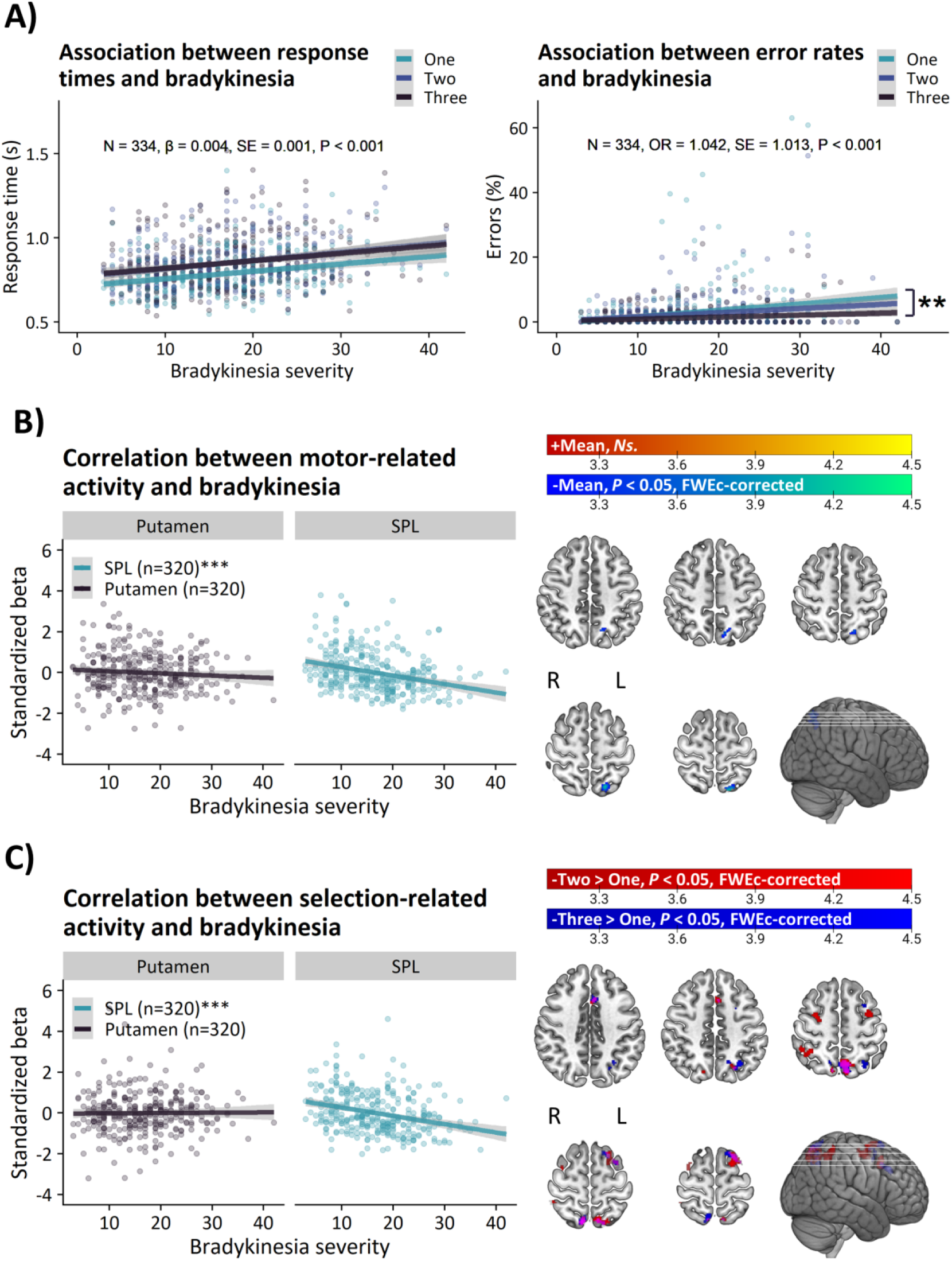
Associations between brain activity and bradykinesia. (**A**) Task performance predicts bradykinesia severity. Greater (**B**) motor-related activity and (**C**) selection-related activity in the superior parietal lobule is associated with lower bradykinesia severity. **=*P*<0.01, ***=*P*<0.001, s=Seconds, SPL=Superior parietal lobule, +/-=Positive/negative correlation. *Ns.*=Not significant, FWEc=Family-wise error cluster. Line-plots show linear associations (solid line) and standard errors (grey area). Brain images show *T*-values of significant clusters.

The influence of action selection demand on error rates depended on bradykinesia severity (Fig. 4A; SEVERITY×CHOICE [*χ*^2^(2)=9.4, *P*=0.009]). That is, patients with more severe bradykinesia had *increased* error rates during low versus high action selection demands (one-choice>three-choice [*β*=0.038, *SE*=0.014, *Z-ratio*=2.7, *P*=0.016]).

There was no effect of SEVERITY on response variability (Supp. Fig. 2E; *P*=0.13) or switching (Supp. Fig. 2F; *P*=0.74).

#### Brain activity

**Motor-related activity:** Lower bradykinesia severity was associated with *greater* motor-related activation of the left superior parietal lobule (VIP; Table 3; Fig. 4B; negative correlation, mean activity>baseline).

**Table 3.** Statistically significant clusters from voxel-wise correlations with bradykinesia severity

| Anatomical label<br>(% cluster volume<br>in area) | Area | P-value<br>(FWEc-<br>corrected) | Cluster extent<br>(voxels) | Max t-value | MNI:<br>X, Y, Z |
| --- | --- | --- | --- | --- | --- |
| **Mean > Baseline** |  |  |  |  |  |
| **Positive correlation** |  |  |  |  |  |
| Ns. |  |  |  |  |  |
| **Negative correlation** |  |  |  |  |  |
| L SPL (76%) | VIP | 0.006 | 144 | 4.2 | -19,-65,60 |
| **Two > One** |  |  |  |  |  |
| **Positive correlation** |  |  |  |  |  |
| Ns. |  |  |  |  |  |
| **Negative correlation** |  |  |  |  |  |
| L SPL (36%) | 7Am | <0.001 | 458 | 4.5 | -11,-63,52 |
| R IPS (78%) | PFm | 0.002 | 161 | 4.5 | 35,-47,52 |
| R SPL (41%) | 7PI | 0.007 | 128 | 4.5 | 11,-67,60 |
| L MFG (46%) | 6ma | <0.001 | 204 | 4.1 | -33,-3,58 |
| R SFG (20%) | 6a | 0.017 | 107 | 4.1 | 29,-5,54 |
| L PCG (55%) | p32pr | 0.049 | 85 | 3.7 | -5,15,46 |
| **Three > One** |  |  |  |  |  |
| **Positive correlation** |  |  |  |  |  |
| Ns. |  |  |  |  |  |
| **Negative correlation** |  |  |  |  |  |
| R SPL (51%) | 7Am | 0.011 | 119 | 4.6 | 9,-65,64 |
| L SPL (39%) | 7Am | <0.001 | 286 | 4.4 | -9,-71,54 |
| L IPS (41%) | MIP | 0.005 | 140 | 4.1 | -29,-63,46 |
| L SFG (47%) | i6-8 | 0.003 | 148 | 4.0 | -15,15,68 |
| L PCG (53%) | p32pr | 0.009 | 123 | 4.0 | -5,17,40 |

Ns.=No significant clusters; SPL=Superior parietal lobule; IPL=Inferior parietal lobule; IPS=Intraparietal sulcus; MFG=Middle frontal gyrus; SFG=Superior frontal gyrus; PCG=Paracingulate gyrus; FWEc=Family-wise error cluster. Anatomical labels were derived from the Anatomy Toolbox v3.0. Area labels were derived from the Glasser atlas.

**Selection-related activity:** At intermediate demands, lower bradykinesia severity was associated with *greater* selection-related activity in the left superior parietal lobule (7Am), right inferior parietal sulcus (PFm), right superior parietal lobule (7Al), left middle frontal gyrus (6ma), right superior frontal gyrus (6a), and left paracingulate gyrus (p32pr; Table 3; Fig. 4C; negative correlation, two-choice>one-choice). At high demands, lower bradykinesia severity was associated with *greater* selection-related activity in the right superior parietal lobule (7Am), left superior parietal lobule (7Am), left intraparietal sulcus (MIP), left superior frontal gyrus (i6-8), and left paracingulate gyrus (p32pr; Table 3; Fig. 4C; negative correlation, three-choice>one-choice).

### The influence of medication

Symptom severity decreased following dopaminergic medication (total: [*χ*^2^(1)=221.1, *P*<0.001, *η*^2^ =0.40]; bradykinesia [*χ*^2^(1)=127.3, *P*<0.001, *η*^2^ =0.29]). There were no effects of dopaminergic medication on task performance or brain activity (see Appendix 3).

## Discussion

We investigated the cerebral mechanisms underlying clinical heterogeneity in Parkinson’s disease in a cohort of early-to-moderately affected^50^ patients (*N*=353) and healthy controls (*N*=60). By leveraging clinical subtyping and brain-symptom associations, we showed that lower symptom severity was consistently associated with higher activation in superior parietal and premotor cortex, particularly during high demands on action selection. In contrast, we found no evidence for a relationship between symptom severity and basal ganglia activity, which was reduced in all clinical subtypes, independently of symptom severity, compared to controls. These findings support the hypothesis that inter-individual variability in symptom severity in Parkinson’s disease may be determined by compensatory cortical processes rather than basal ganglia dysfunction.

### Action selection deficits are accompanied by basal ganglia dysfunction and bradykinesia

Consistent with previous neuroimaging and behavioural studies, we found that Parkinson’s disease was associated with reduced motor-related activity in a core network of sensorimotor regions (putamen, primary motor cortex, and cerebellum), and this was accompanied by general slowing of motor performance and increased error rates.^13, 14, 64, 65^ Furthermore, we observed that slower responses and increased error rates were both associated with higher severity of bradykinesia. In combination, these findings suggest that our task was sensitive to action selection deficits and basal ganglia dysfunction.

This sensitivity was built into this task by virtue of two design improvements over previous action selection tasks.^18, 47, 66^ First, we varied the number of response options parametrically on a trial-by-trial basis, using a mixed block/event-related design with a jittered inter-stimulus interval.^56–58^ We also explicitly instructed participants to make equal use of all response options. This reduced predictability in the task, motivating participants to generate responses based on a process of selection rather than a pre-defined strategy, such as responding with the same finger on each trial, which may be more common in patients than controls.^6^ Indeed, we found no between-group differences in response variability or switching, indicating that all groups utilized the same behavioural strategy. The parametric variation of response options additionally enabled us to investigate the effects of increasing demands on action selection. Our results indicate that compensatory cerebral mechanisms are recruited when demands increase (see below). Second, patients were asked to respond with their most affected hand, which enhances sensitivity to Parkinson’s disease-related deficits and excludes a potential difference in action selection capacity between the right and left sides.

### Evidence for a compensatory role of parieto-premotor cortex

Our study provides evidence that the parieto-premotor cortex may support a compensatory role in Parkinson’s disease. Conceptually, compensatory cerebral alterations involve a performance-enhancing recruitment of neural resources that enable patients to carry out movements despite deficits in the neural mechanisms that typically support the performance of those movements.^30, 35, 36^ Accordingly, motor impairments are expected to emerge when dysfunction exceeds the capacities of compensatory mechanisms that support motor performance, such as increased reliance on sensory cueing^67^ or goal-directed control.^6^ This may explain why motor symptoms differentially worsen during voluntary movements that require a selection between multiple actions.^15–18^

Our findings fit with a compensatory role for the parieto-premotor cortex in Parkinson’s disease, for the three following reasons. First, we demonstrate that Parkinson’s disease is associated with decreased basal ganglia activity and impaired action selection performance. This demonstrates the presence of dysfunction in one brain region, which would in turn call for compensation elsewhere in the nervous system. Second, we report upregulated action selected-related activity in the parieto-premotor cortex of Parkinson patients with the most benign clinical subtype. Specifically, compared to healthy controls, mild-motor predominant patients showed *increased* parieto-premotor activity, whereas diffuse-malignant patients showed *reduced* premotor activity. The lack of between-group differences in response variability and switching further suggests that these effects were not caused by differences in the ability to perform the task. Third, we provide evidence that upregulated parieto-premotor activity is not just a by-product of inefficient cerebral processing: across the entire cohort, stronger enhancement of parieto-premotor activity was associated with lower severity of bradykinesia symptoms. In combination, these findings suggest that enhanced parieto-premotor function may confer beneficial effects on motor performance by partially compensating for deficits in basal ganglia function,^30, 68^ which fits with the recent reconceptualization of bradykinesia as a consequence of large-scale network dysfunction.^2, 8, 69–71^ Accordingly, we predict that the worsening of motor symptoms is primarily driven by decline in parieto-premotor compensation (Fig. 5A). We are currently addressing this hypothesis in an upcoming study where we extend the methodology of the present study to a longitudinal setting.

**Fig. 5.**
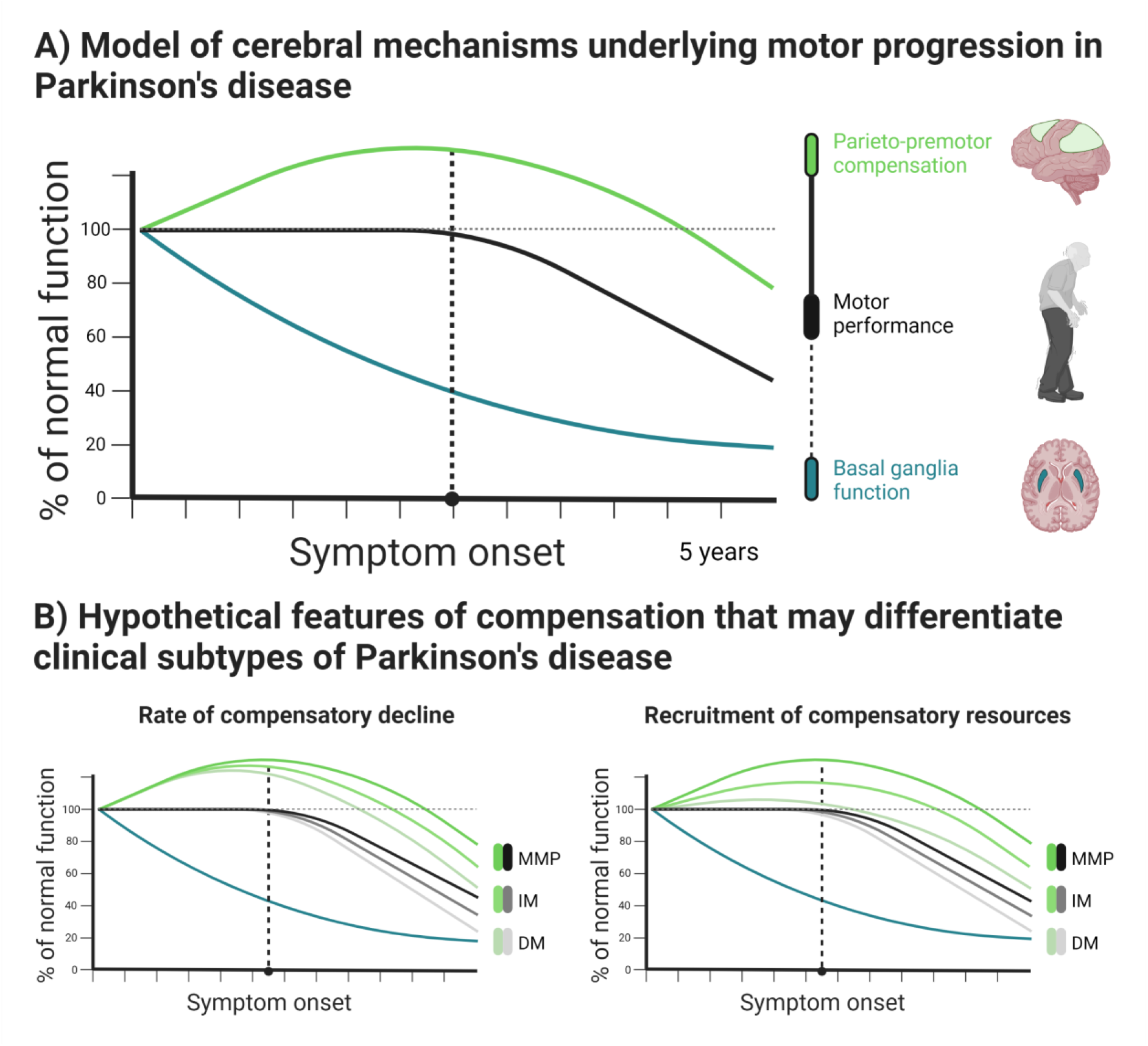
Model of the relationship between changes in brain activity and motor symptom progression. (**A**) Parieto-premotor activity is enhanced to compensate for basal ganglia dysfunction during the pre-symptomatic phase of Parkinson’s disease. Eventually, parieto-premotor compensation begins to decline, leading to the emergence of motor impairments caused by basal ganglia dysfunction. According to this model, decline in motor performance depends on loss of cortical compensation rather than progressive basal ganglia dysfunction. (**B**) Subtypes may be differentiated by the rate at which cortical compensation declines, the degree to which cortical resources can be recruited for compensatory purposes, or a combination of the two.

### Neural mechanisms underlying focal and diffuse clinical subtypes

Subtypes of Parkinson’s disease have been linked to functional and structural alterations that may contribute to differences in clinical phenotype, symptom severity, and progression.^72^ For example, the diffuse-malignant subtype, which is characterized by relatively severe motor and non-motor symptoms,^73^ is associated with increased excitability and reduced plasticity in the primary motor cortex,^74^ wide-spread disruptions in structural connectivity,^75^ and more extensive atrophy at both cortical and sub-cortical levels.^51, 75^ These alterations may reflect decline in neural reserves and impaired maintenance of neural resources.^30^ We add to these findings by showing that the diffuse-malignant subtype is characterized by more extensive cortical dysfunction, encompassing parietal, premotor, and primary somatosensory cortex, but not by a differential reduction in basal ganglia dysfunction. More specifically, we show that the diffuse-malignant subtype is characterized by deficits in the compensatory upregulation of parieto-premotor activity during action selection, which remains intact in the mild-motor predominant subtype. Weaker compensation in diffuse-malignant patients may result from deficits in the ability to recruit compensatory resources (Fig. 5B, left), faster decline in compensatory function (Fig. 5B, right), or both.

Variability in the recruitment and decline of compensatory function is likely dependent on subtype-specific differences in pathological mechanisms, such as the spread of α-synucleinopathy.^76^ It has been proposed that α-synucleinopathy can begin either in the central nervous system, spreading towards the peripheral nervous system (“brain-first”), or the other way around (“body-first”).^77^ These two propagation modes of α-synucleinopathy have been linked to distinct clinical phenotypes that resemble the subtypes that we utilized in this study.^78^ That is, a brain-first form of α-synucleinopathy (which overlaps with the clinical “mild motor-predominant” subtype) has been linked to younger disease onset and motor symptoms that are confined to single effectors, likely as a result of somatotopically dependent retrograde nigral degeneration.^79^ The body-first form, on the other hand, has been associated with older disease onset, diffuse symptomatology involving both motor and non-motor domains, and rapid clinical progression. Therefore, this form overlaps with the clinical “diffuse-malignant” subtype. These differences in symptomatology suggest that a brain-first type of pathology may lead to focal deficits in brain function that are restricted to cortico-striatal loops whereas a body-first type may lead to diffuse deficits that involve more widespread cortical networks. Hence, the diffuse-malignant subtype may be characterized by weaker parieto-premotor compensation as a result of diffuse cortical α-synucleinopathy, which effectively constrains the neural resources that patients with this subtype are able to recruit for compensatory purposes.

### Mechanisms underlying parieto-premotor compensation

While the mechanisms that enable the parieto-premotor cortex to compensate for basal ganglia dysfunction cannot be directly inferred from this study, we suggest that they may involve non-motor territories of the basal ganglia (anterior striatum and the caudate nucleus)^68, 80, 81^ and motor control pathways that bypass the basal ganglia.^82^ A recent theory holds that sensorimotor regions specify and select between competing motor responses through a process of evidence accumulation towards a decision boundary.^83, 84^ This process is dynamically influenced by urgency signals originating from the basal ganglia that determine the vigour of responses in a contextually dependent manner. In Parkinson’s disease, urgency signalling is disrupted by dopamine depletion, thereby increasing the amount of evidence that sensorimotor regions must accumulate to activate motor patterns, leading to loss of movement vigour (slowness and inability to initiate movements). However, sensorimotor regions receive additional biasing inputs that may be relied on to compensate for basal ganglia dysfunction. For example, cognitive and affective territories of the basal ganglia that are relatively spared from dopamine depletion in earlier stages of Parkinson’s disease may become increasingly involved in action selection.^6, 68, 85^ Furthermore, movements can be triggered via pathways that bypass the basal ganglia,^82^ which is consistent with a wealth of evidence suggesting that patients can rely on sensory input, such as visual cues,^67^ to compensate for movement deficits. We therefore speculate that an upregulation of parieto-premotor function in Parkinson’s disease may reflect increased processing of inputs from non-motor territories of the basal ganglia as well as other cortical regions that enable sensorimotor regions to exceed decision boundaries and select actions despite dysfunctional urgency signalling.

### Limitations and interpretational issues

Previous research has demonstrated a differential effect of action selection on response times in patients with Parkinson’s disease compared to healthy controls.^15–18^ We did not observe such an effect in our study. Instead, we found that Parkinson’s disease was associated with a slowing of response times across levels of action selection demand, suggesting a general deficit in action selection performance. This may relate to the relatively short disease duration of our cohort (maximally 5 years), raising the possibility that deficits in behavioural performance resulting from increased action selection demand mainly arise in later stages of Parkinson’s disease, potentially as a result of decline in compensatory resources. This possibility receives some support from our finding that that action selection led to a greater increase in response times for intermediate patients compared to the mild-motor predominant patients, suggesting that motor deficits may worsen as action selection demand increases. The relatively small sample size of diffuse-malignant patients may explain why we did not find the same effect in this group.

The lack of a differential effect of action selection demand on task performance between Parkinson’s disease patients and healthy controls eliminates the potential confound of different behavioural strategies, thereby improving the interpretability of the between-group effects on brain activity that we observed.^86^ This claim is further substantiated by a lack of between-group effects on response variability and switching, suggesting that patients and healthy controls were equally capable of using all available response options.

Increased parieto-premotor activation could reflect alterations in saccadic eye-movement control rather than the activation of compensatory processes.^87, 88^ Arguing against this interpretation, we found no clusters that centred on typical saccade control regions, such as the frontal and posterior eye fields.

The lack of an effect of medication on parieto-premotor function indicates that cortical compensation in Parkinson’s disease may not be influenced by dopaminergic state, and may therefore require support from alternative therapeutic approaches.^5^ However, despite clear improvements in motor symptoms following dopaminergic medication, we did not observe normalization of response times^89^ or basal ganglia function.^13^ One possible explanation is that patients were not completely OFF, due to long-lasting medication effects which are now well recognised.^90^ The effect of levodopa on motor symptoms is mediated by a short-duration response that alleviates symptoms within hours and a long-duration response that provides symptomatic relief over days to weeks.^91^ The long-duration response is particularly strong in early-stage Parkinson’s disease, meaning that the clinically defined off-state in which participants were assessed, defined as the withdrawal of dopaminergic medication for at least 12 hours, may not have allowed the effect of medication to subside enough to detect subtle alterations in task performance and brain activity.^90^

Parkinson’s disease is associated with wide-spread brain atrophy that may have driven inter-individual differences in compensatory capacity in this study.^76, 92, 93^ Contrary to these findings, we observed no grey matter volume alterations in parieto-premotor areas where differences in selection-related activity between clinical subtypes of Parkinson’s disease and healthy controls were located (see Appendix 4). Instead, differences in grey matter volume were confined to a more inferiorly located network consisting of occipital, inferior parietal, and temporal areas of the cortex, which is consistent with previous findings of brain atrophy in early-to-moderate stages of Parkinson’s disease (see Supp. Fig. 6).^93^ We therefore conclude that compensatory alterations in parieto-premotor cortex primarily reflect functional neuroplasticity rather than alterations in underlying brain structure.

## Conclusion

Our results suggest that inter-individual variability in Parkinson’s disease with respect to clinical phenotype and bradykinesia severity are determined, in part, by the degree to which parieto-premotor cortex is able to compensate for progressive basal ganglia dysfunction. Interventions and treatments that aim to modify the progression of Parkinson’s disease may benefit from focussing on enhancing the efficiency and maintenance of compensatory cortical processes in addition to restoring basal ganglia dysfunction.

## Acknowledgements

We would like to extend a sincere thank you to everyone who participated in the Personalized Parkinson Project. We would also like to thank Paul Gaalman and Jose Marques for their assistance with the MRI protocol that was used in this study, and Freek Nieuwhof for his valuable assistance during the initiation of this study.

## Supplementary material

Supplementary material is available at *Brain* online.

## Abbreviations

MDS-UPDRS: Movement Disorders Society Unified Parkinson Disease Rating Scale

### Appendix 1 Inclusion and exclusion criteria

Patients were eligible for participation if they had received a diagnosis of idiopathic Parkinson’s disease from a certified neurologist, had 0-5 years disease duration, and were ≥ 18 years of age. It should be noted that the Personalized Parkinson Project cohort did not constitute a convenience sample, but rather aimed to include a cohort that represented real-life patients. Strict stratification criteria were applied to ensure a balanced inclusion of men and women, different age ranges (21-45; 46-55; 56-65; ≥66 years), and different disease durations (<2.5 years; ≥2.5 years). Healthy controls were eligible for participation if they were at least 40 years of age and were willing and able to return for a two-year follow-up measurement. All participants were able to read and understand Dutch, could comply with all aspects of the study protocol, and could provide written informed consent. Exclusion criteria included co-morbidities that would negatively influence the interpretability of parkinsonian disability, contraindications to MRI, pregnancy or breastfeeding, and nickel allergy (owing to the wearing of a study-related device). Additional exclusion criteria for healthy controls included co-morbidities that would negatively influence the interpretability of results from a comparison with PD patients. Further details can be found in the primary study protocol of the Personalized Parkinson Project.^1^

#### Measurements used for subtype classification

Motor symptoms were assessed with a composite score based on MDS-UPDRS II and III.^2^ Cognitive function was assessed with a composite score derived from age-, education-, and sex-adjusted z-scores on the Benton Judgement of Line Orientation,^3^ Brixton Spatial Anticipation Test,^4^ Semantic Fluency Test (1 minute animal naming),^5^ Symbol Digit Modalities Test (90 seconds, oral version),^6^ Letter-Number Sequencing from the Wechsler Adult Intelligence Test – Fourth Edition,^7^ and an average across subscores of the Rey Auditory Verbal Learning Test.^5, 8^ REM sleep behavior disorder was assessed with the REM Sleep Behavior Disorder Screening Questionnaire.^9^ Autonomic function was assessed with the Scales for Outcomes in Parkinson’s disease.^10^

#### Detailed description of the action selection task

Each trial began with the presentation of a fixation cross. After a random inter-stimulus interval of 2-4 seconds, the cross was replaced by a cue consisting of four circles, with each circle corresponding to a button on a response device. The circles were either filled or empty to indicate which responses were correct or incorrect, respectively. Participants were instructed to respond to cues by pressing a single button corresponding to a single filled circle. If multiple circles were highlighted, then participants were instructed to choose which circle to respond to. Only one response was allowed per trial. Each cue included either one, two, or three filled circles, thereby varying the number of response choices that participants were presented with on a trial-by-trial basis. Participants were encouraged to make use of all four buttons rather than selectively responding with only a subset of buttons, to vary the finger that was used to respond during trials where multiple response options were presented, and to respond as quickly and as accurately as possible. Cues remained on the screen for a maximum of 2 seconds or until a response was recorded and were immediately followed by the presentation of a new fixation cross. The task consisted of 132 trials and lasted for approximately 10 minutes, depending on performance. There were 60 one-choice trials (15 per finger), 30 two-choice trials, and 30 three-choice trials. The additional 12 trials consisted of 6 one-choice trials, 3 two-choice trials, and 3 three-choice trials where circles were outlined in red. Participants were instructed to withhold a response during these “catch” trials. Trials were presented in three blocks, each consisting of 44 trials. Out of these 44 trials, 20 were one-choice, 10 were two-choice, 10 were three-choice, and 4 were catch. The ordering of trial conditions within each block was pseudo-randomized. Blocks were separated by 20 seconds of rest. Prior to entering the scanner, participants practiced by performing one continuous block of 68 trials (30 one-choice trials, 15 two-choice trials, 15-three choice trials, 8 catch trials). Patients were asked to perform the task with their most-affected hand. The responding hands of healthy controls were matched to the 56 patients who were assessed in an off-medicated state.

#### Preprocessing details

Linear transformations of functional images to anatomical space were estimated using a linear transformation with boundary-based registration and six degrees-of-freedom.^11^ Non-linear transformations were estimated from anatomical to MNI152Lin6Asym-space.^12, 13^ A single interpolation step was used to carry out all transformations in combination with motion correction^14^ and slice time correction.^15^ Confound time series were generated for framewise displacement and DVARS,^16^ along with 24 motion derivatives. An anatomical principal component analysis was performed to derive time series for cerebrospinal fluid and white matter signal.^17^ ICA-AROMA was used to derive time series of motion-related noise^18, 19^ whose classification was further refined using custom methodology (see section below). A series of discrete cosine-basis functions were derived for high-pass filtering (>0.008 Hz). Tremor regressors were generated from the accelerometry data of 69 patients.

#### Refinement of ICA-AROMA component selection

Additional steps were taken to ensure that the removal of confounding motion-related variability through ICA-AROMA during the first-level analysis did not adversely affect the estimation of task-related regressors. For each participant, the classification of ICA-AROMA components was refined in a multiple regression analysis where each noise time series was modelled as a function of task regressors for choice and catch conditions. Time series that shared more than 5% explained variance, as assessed by the *r*^2^ of the regression model, were reclassified as non-noise and subsequently left out of the first-level design.

#### Quantification of tremor

For all patients, tremor severity was quantified using a three-axial accelerometer placed on the dorsum of the most-affected hand. Accelerometry preprocessing involved detrending, demeaning, transformation to scan-to-scan tremor power at peak frequency, and log-transformation, after which the tremor signal was convolved with a canonical hemodynamic response function.^20^ The resulting tremor regressors were added to the first-level models of 69 patients whose tremor was confirmed through visual inspection.

### Appendix 2 Task-related activity across groups

Task-related activity was investigated across groups to generate activation maps of motor-activity, catch-related activity, intermediate selection-related activity, and high action selection-related activity.

#### Motor-related activity

A conjunction analysis of one-choice, two-choice, and three-choice activity yielded a network of cerebellar, visual, parietal, insular, and sensorimotor activity (Supp. Fig. 1A). Sensorimotor activity was more extensive in the left hemisphere, as would be expected given that this was the responding side of all participants, either naturally or as a result of horizontal flipping of contrast images.

#### Catch-related activity

Catch-related activity primarily captured visual, insular, and prefrontal activity (Supp. Fig. 1B).

#### Action selection-related activity

Intermediate (Supp. Fig. 1C) and high (Supp. Fig. 1D) demand on action selection elicited increased activity in a frontoparietal network and decreased activity in regions involved in the default mode network. This is consistent with the idea that action selection led to the recruitment of additional cognitive processing beyond simple motor responses.

**Supp. Fig. 1.**
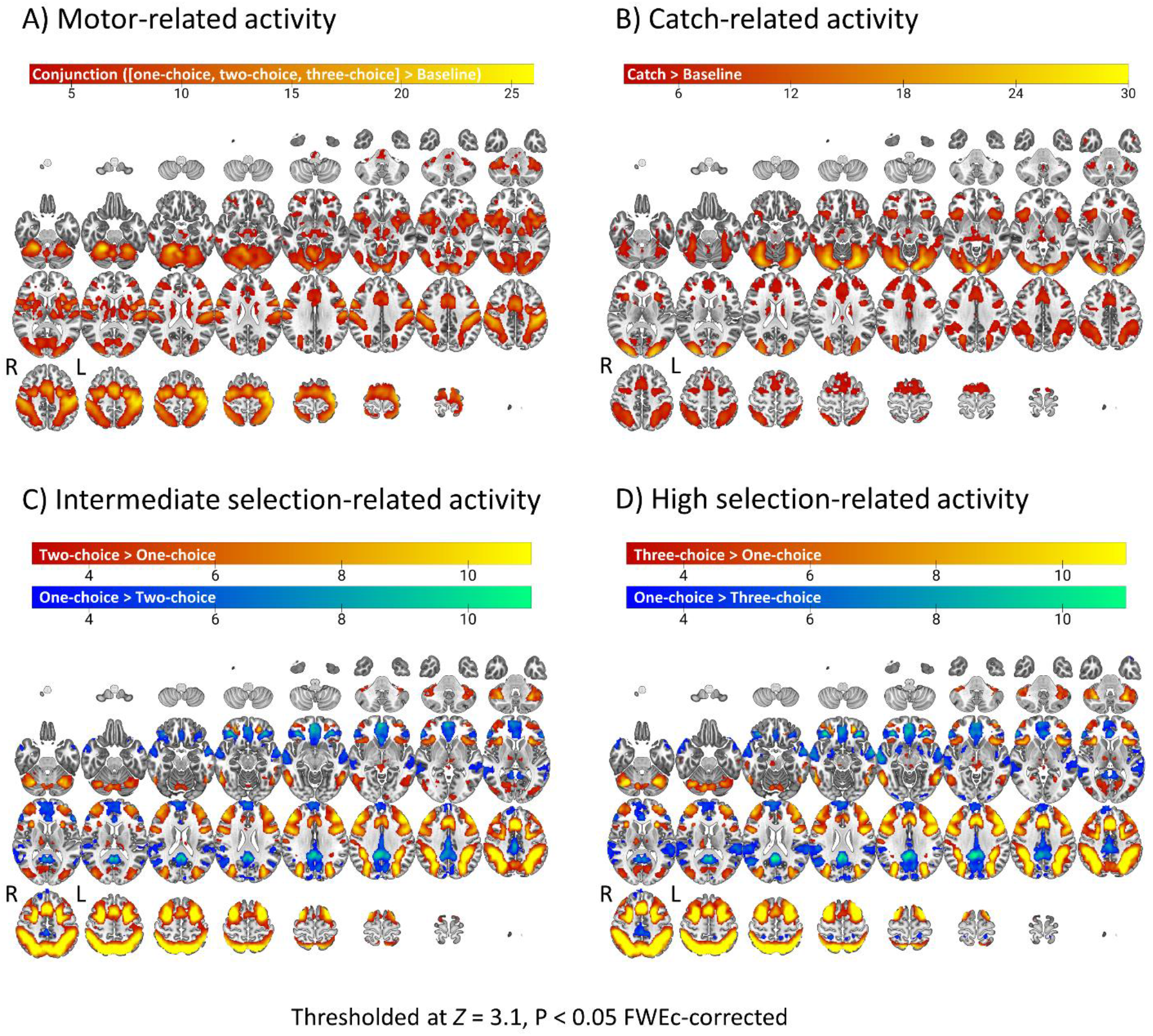
Task-related activity across patients and healthy controls. (A) Sensorimotor network activation common to all levels of action selection demand. (B) Response withholding preferentially activates visual and prefrontal cortex. (C) Intermediate and (D) high action selection demand leads to activation of the frontoparietal network and deactivation of the default mode network.

**Supp. Fig. 2.**
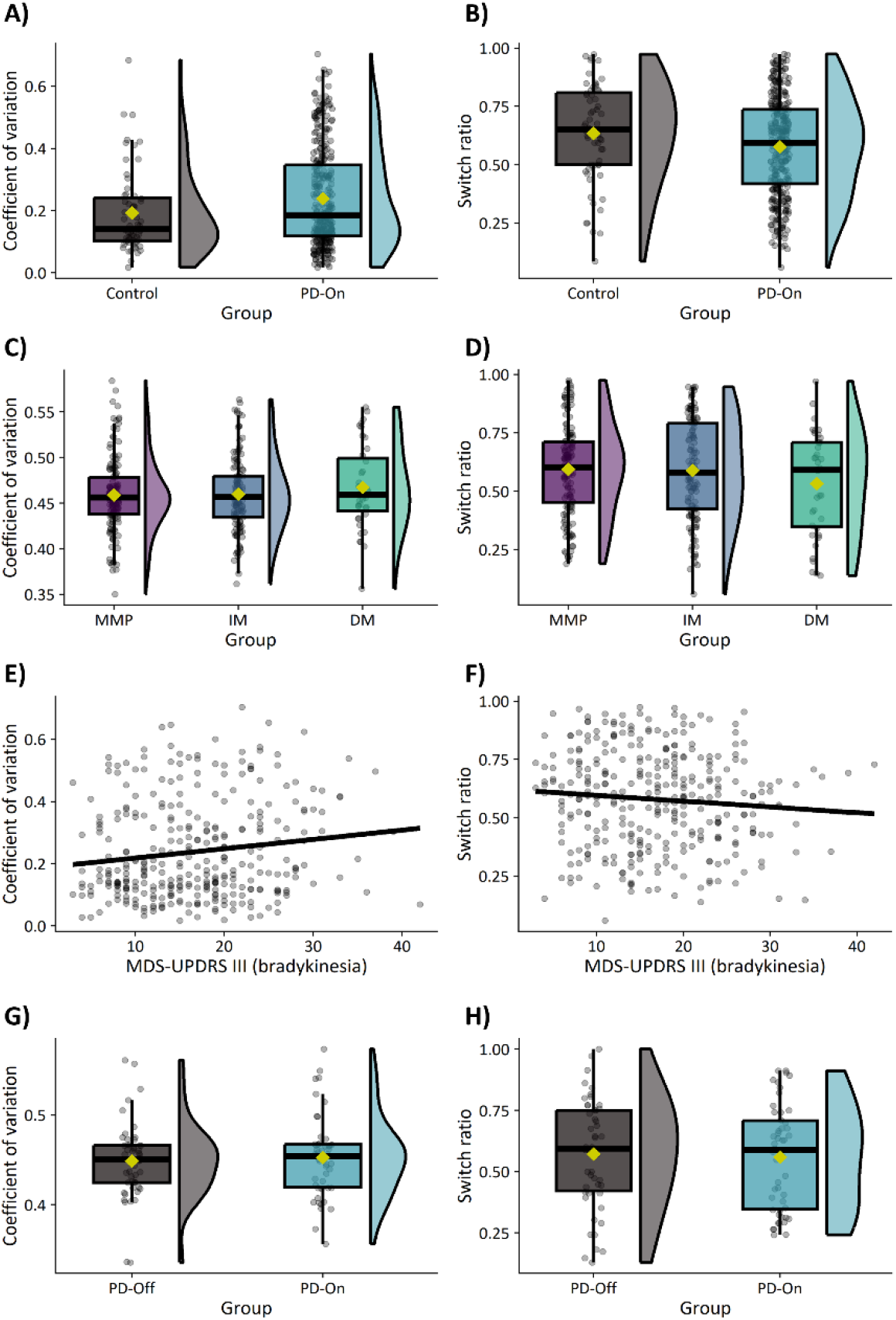
Effects on response variability (left) and switching (right). (A/B) Effect of Parkinson’s disease. (C/D) Effect of subtype. (E/F) Associations with bradykinesia. (G/H) Effect of medication.

**Supp. Fig. 3.**
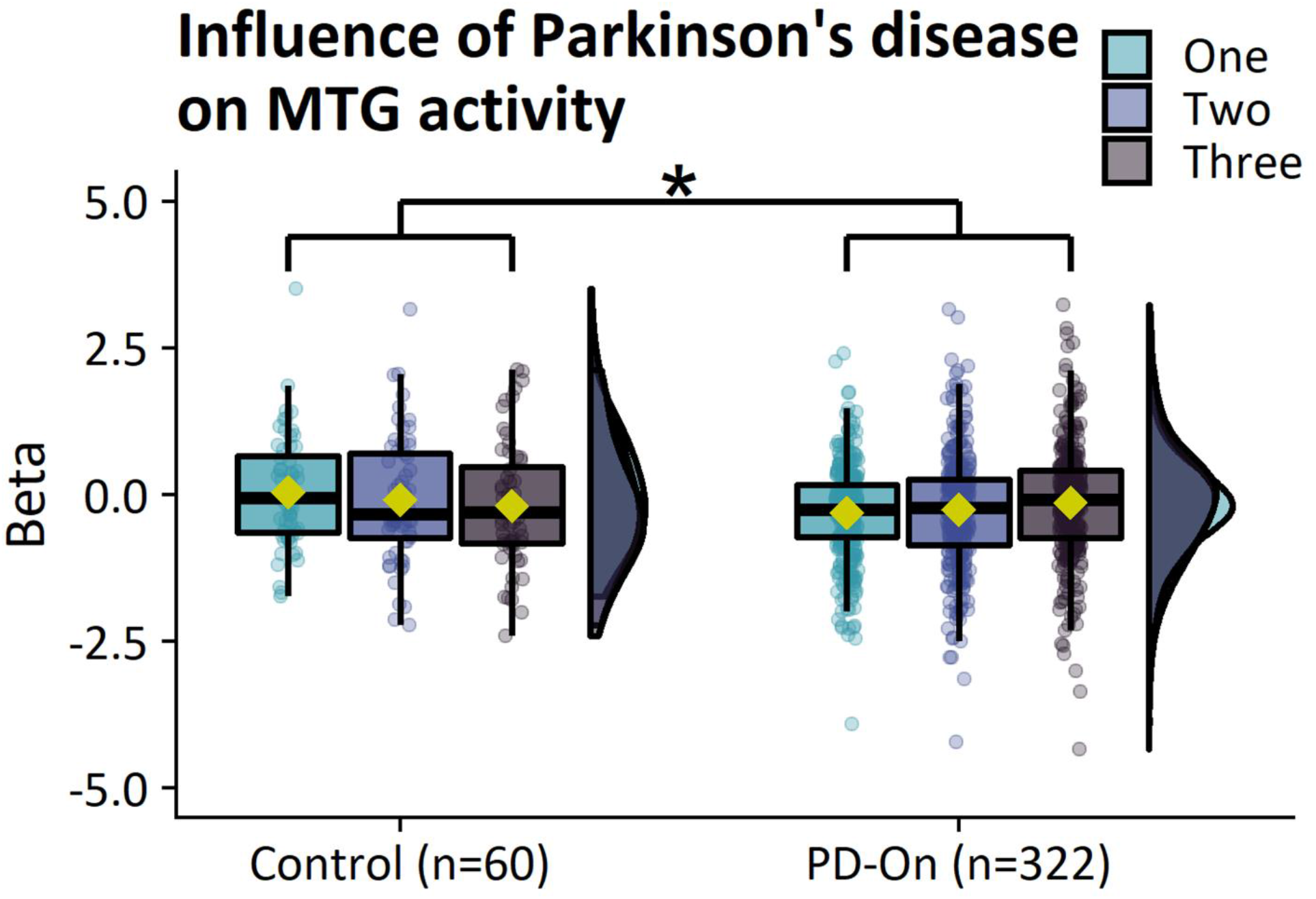
Selection-related deactivation of the inferior parietal lobule. Patients show reduced deactivation of the inferior parietal lobule in comparison to healthy controls.

**Supp. Fig. 4.**
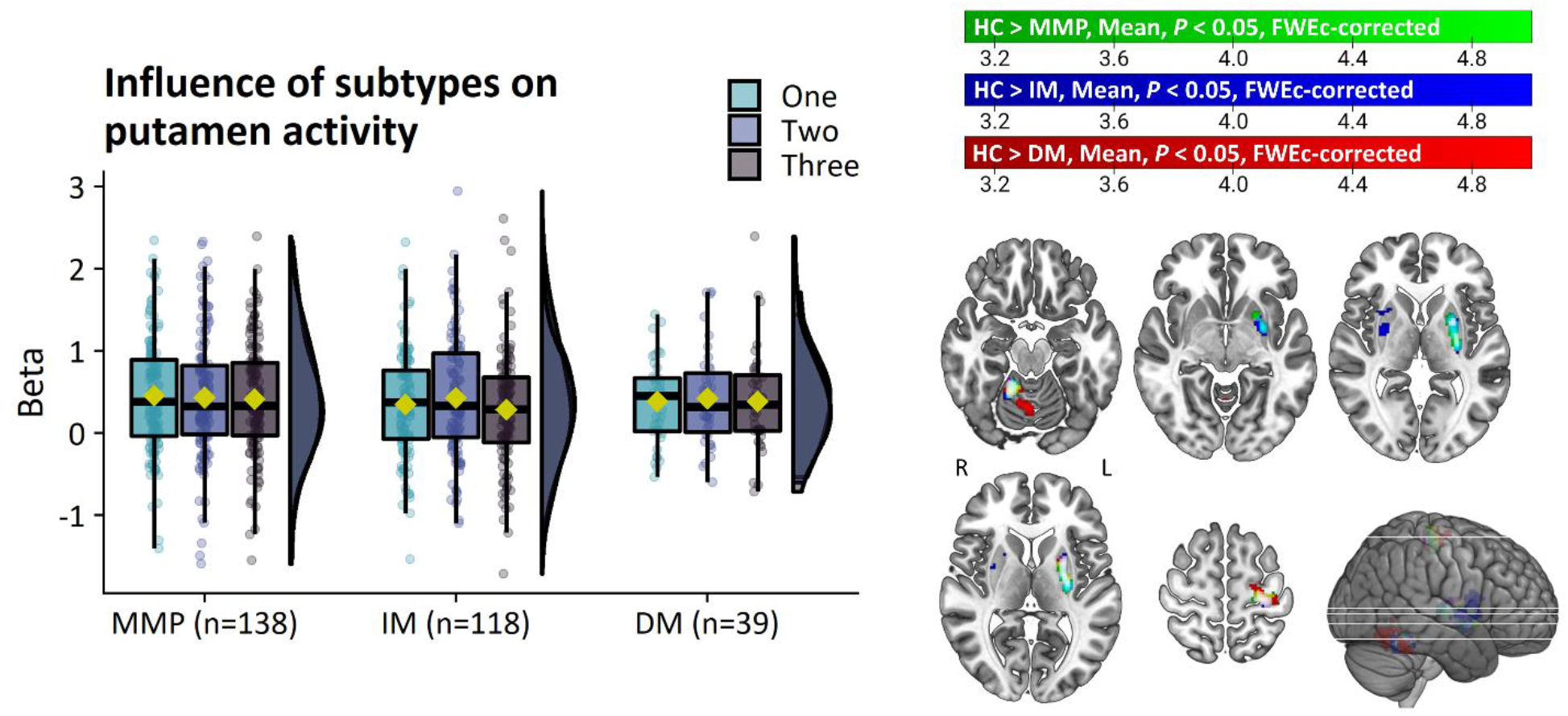
Similar deficits in basal ganglia function between subtypes. Subtypes show no differences in putamen activity in the most affected hemisphere (right). In comparisons to healthy controls, all subtypes show decreased motor-related activity in a common network of the putamen, primary somatosensory cortex, and cerebellum (left).

### Appendix 3 The influence of medication

#### Behavioral performance

Response times increased as a function of action selection demand (Supp. Fig. 4; main effect of CHOICE [*χ*^2^(2)=22.9, *P*<0.001, *η^2^_p_*=0.07]; intermediate>low [*log-ratio*=1.06, *SE*=0.009, *t-ratio*(843)=7.0, *P*<0.001], high>low [*log-ratio*=1.06, *SE*=0.009, *t-ratio*(843)=6.4, *P*<0.001]). There was no effect of medication on response times (*P*=0.36).

Error rates increased as a function of action selection demand (Supp. Fig. 4; main effect of CHOICE [*χ*^2^(2)=9.5, p=0.009]; low>high [*OR*=2.5, *SE*=0.53, *Z-ratio*=4.4, *P*<0.001], intermediate>high [*OR*=2.2, *SE*=0.52, *Z-ratio*=3.4, *P*=0.002]). There was no effect of medication on error rates (*P*=0.56).

There was no effect of medication on response perseverance (Supp. Fig. 2G; *P*=0.72) or switching (Supp. Fig. 2H; *P*=0.97).

#### Brain activity

**Motor-related activity:** There were no effects of medication on motor-related activity.

**Selection-related activity:** There were no effects of medication on selection-related activity.

**Supp. Fig. 5.**
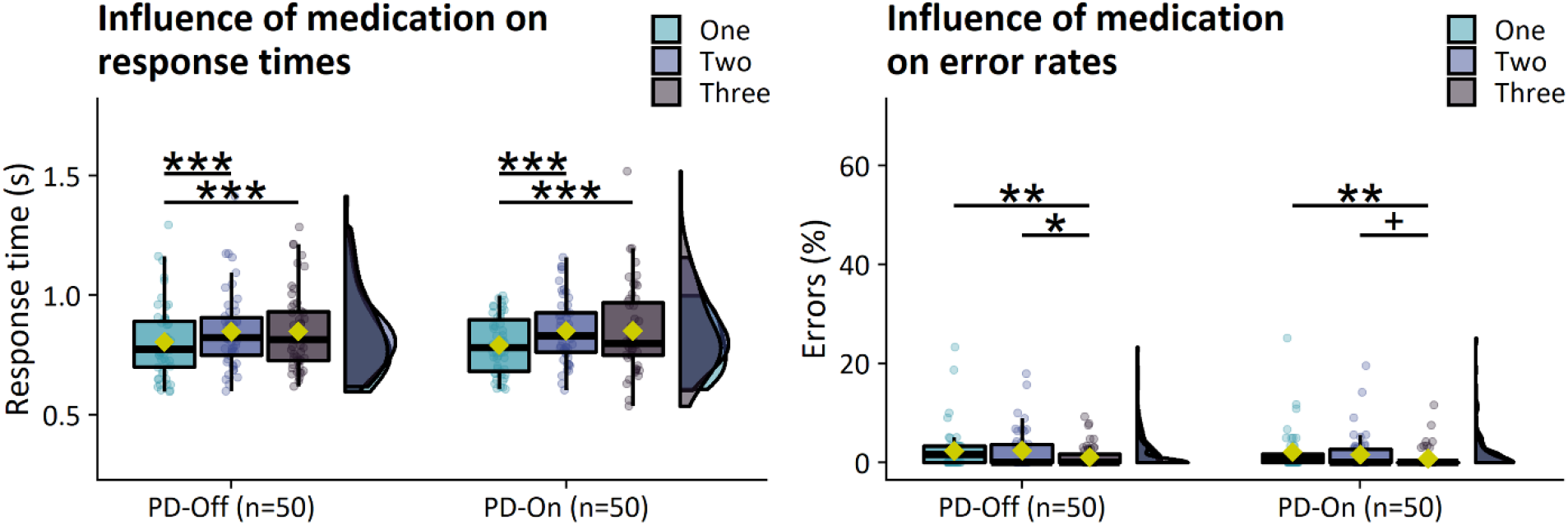
Influence of medication on response times (left) and error rates (right).

### Appendix 4 Voxel-based morphometry

#### Processing and statistical analysis of gray matter volume in regions showing group effects on brain activity

Functional alterations in Parkinson’s disease may be associated with underlying patterns of structural atrophy. Voxel-based morphometry (VBM)^21^ was used to estimate gray matter volume in brain regions that showed significant differences in brain activity between (1) patients and controls, (2) subtypes, and (3) subtypes and controls. The Computational Anatomy Toolbox^22^ (CAT; version 12.8) was used to segment T1-weighted structural images into gray matter, white matter, and cerebrospinal fluid. Gray matter segmentations were normalized to a study-specific gray matter template that was generated with geodesic shooting nonlinear image registration^23^ based on 59 healthy controls and 59 randomly selected PD patients. The normalized gray matter images were subsequently modulated by Jacobian determinants to account for the non-linear component of the normalization procedure.

Average gray matter volumes were separately extracted from networks consisting of clusters that showed a significant effect of GROUP on brain activity. Volumes were extracted from contralateral networks for participants whose first-level contrast had been flipped to ensure correspondence between functional and structural analyses. The grand-average gray matter volume of each network was calculated per participant, yielding a single metric of gray matter volume per group comparison. One-way ANCOVAs were used to test the effect of GROUP on gray matter volume within each network. Total intracranial volume and age were modelled as covariates of non-interest. Comparisons that yielded significant between-group differences were followed with an exploratory analysis of the association between gray matter volume and bradykinesia severity in all patients with Parkinson’s disease. Whole-brain voxel-wise comparisons were conducted to explore more extensive differences in gray matter volumes.

Voxel-wise whole-brain comparisons revealed decreased gray matter volume in patients compared to controls in a network consisting of large areas of visual, temporal, orbitofrontal, posterior parietal cortex, and the anterior striatum (Supp. Fig. 4). Parietal cortex atrophy was primarily lateralized to the left hemisphere.

#### Group comparisons of gray matter volume

##### Patients versus controls

The region of the middle temporal cortex where patients showed reduced selection-related deactivation compared to controls (patient>control, three-choice>one-choice) also showed decreased gray matter volume (control>patient [*F*(1)=8.9, *P*=0.003, *η^2^_p_*=0.02]). Lower gray matter volume in this region was associated with higher bradykinesia severity *(β*=-0018, *95% CI* = [−0.0004, −0.0032], *T*(301)=2.5, *P*=0.012).

##### Subtypes

The region of the inferior parietal lobule (area PFt) where the diffuse-malignant subtype showed decreased motor-related activity compared to the mild-motor predominant subtype (mild-motor predominant>diffuse-malignant, mean>baseline) also showed a decrease in gray matter volume (mild-motor predominant>diffuse-malignant [*F*(1)=6.8, *P*=0.011, *η*^2^ =0.05]). Lower gray matter volume in this region was associated with higher bradykinesia severity (*β*=-0023, *95% CI*=[−0.0007, −0.0040], *T*(301)=2.8, *P*=0.006).

The network where the mild-motor predominant subtype showed increased selection-related activity compared to controls (mild-motor predominant>control, three-choice>one-choice) showed a decrease in gray matter volume (control>mild-motor predominant [*F*(1)=8.7, *P*=0.004, *η^2^_p_*=0.05]). Lower gray matter volume in this network was associated with higher bradykinesia severity (*β*=-0012, *95% CI*=[−0.0002, −0.0022], *T*(301)=2.3, *P*=0.021). A post hoc analysis of each separate region of the network showed that this effect was driven primarily by the region of middle temporal cortex (control>mild-motor predominant [*F*(1)=8.1, *P*=0.005, *η^2^_p_*=0.05]) where patients showed an overall decrease compared to controls. There were no significant differences between subtypes at the voxel-wise whole-brain level.

**Supp. Fig. 6.**
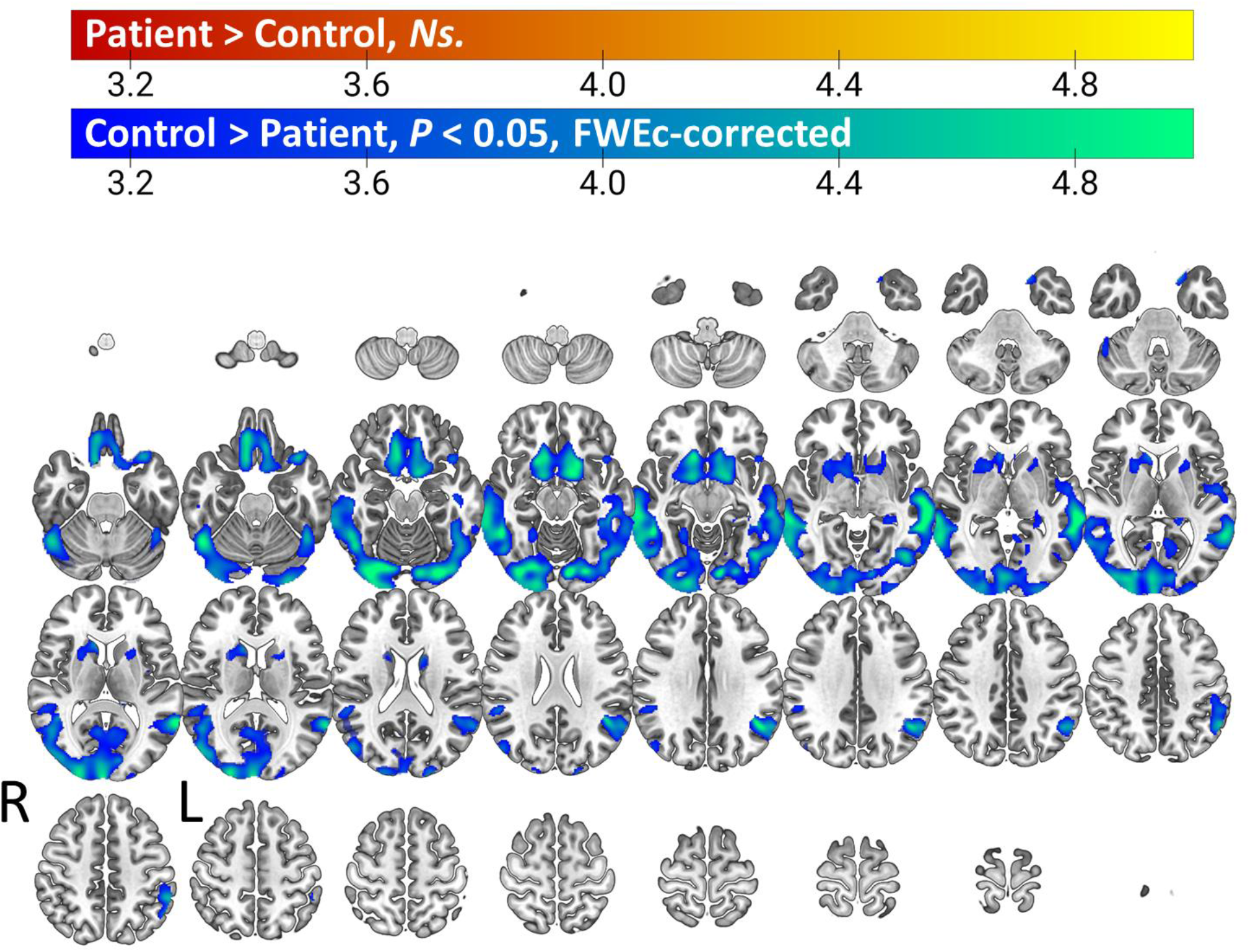
Gray matter volume reductions in Parkinson’s disease. Voxel-wise comparison of gray matter volume between patients and healthy controls reveals a network of atrophy consisting of large regions of visual, temporal, inferior parietal, and orbitofrontal cortex, as well as anterior regions of the striatum.

